# HPVsim: An agent-based model of HPV transmission and cervical cancer

**DOI:** 10.1101/2023.02.01.23285356

**Authors:** Robyn M. Stuart, Jamie A. Cohen, Cliff C. Kerr, Prashant Mathur, National Disease Modeling Consortium of India, Romesh G. Abeysuriya, Marita Zimmermann, Darcy W. Rao, Mariah C. Boudreau, Serin Lee, Luojun Yang, Daniel J. Klein

## Abstract

In 2020, the WHO launched its first Global Strategy to Accelerate the Elimination of Cervical Cancer, outlining an ambitious set of targets for countries to achieve over the next decade. At the same time, new tools, technologies, and strategies are in the pipeline that may improve screening performance, expand the reach of prophylactic vaccines, and prevent the acquisition, persistence and progression of oncogenic HPV. Detailed mechanistic modeling can help identify the combinations of current and future strategies to combat cervical cancer. Open-source modeling tools are needed to shift the capacity for such evaluations in-country. Here, we introduce the Human papillomavirus simulator (HPVsim), a new open-source software package for creating flexible agent-based models parameterized with country-specific vital dynamics, structured sexual networks, and co-transmitting HPV genotypes. HPVsim includes a novel methodology for modeling cervical disease progression, designed to be readily adaptable to new forms of screening. The software itself is implemented in Python, has built-in tools for simulating commonly-used interventions, includes a comprehensive set of tests and documentation, and runs quickly (seconds to minutes) on a laptop. Performance is greatly enhanced by HPVsim’s multi-scale modeling functionality. HPVsim is open source under the MIT License and available via both the Python Package Index (via pip install) and GitHub (hpvsim.org).

## 1. Introduction

In August of 2020, the World Health Organization adopted its first ever strategy to permanently end cervical cancer as a global public health problem (1). The Global Strategy to Accelerate the Elimination of Cervical Cancer proposes targets of 90% of girls vaccinated against human papillomavirus (HPV, the virus that causes cervical cancer) by age 15, 70% of women screened by age 35 and again by 45, and 90% compliance with treatment recommendations for pre-cancer and management of invasive cancer. The ability for countries to achieve these ambitious targets was bolstered in April of 2022, when the WHO Strategic Advisory Group of Experts on Immunization (SAGE) completed a review of HPV vaccine dosing schedules and concluded that one-dose of the vaccine provides comparable efficacy to two or three-doses (2). With more doses now available, the pathway towards cervical cancer elimination has become more achievable for many countries, including those with the highest burden.

The formulation of the goal to eliminate cervical cancer and the strategies that might get us there has been informed by the results of a well-established set of mathematical models (3–8). Models have also been instrumental in evaluating and incorporating new technologies into guidelines and health policies worldwide (9–13). While these models continue to provide valuable insight to guide cervical cancer elimination, there are nevertheless modeling questions that call for a different methodological approach. Firstly, shifts in our understanding of HPV epidemiology and the emergence of new biomarker-based screening tools have prompted calls for a new generation of cervical cancer models that simulate the underlying biological processes in addition to their clinical correlates (14). Secondly, demand for open-source software in general, and open-source scientific models in particular, has been continually increasing in response to tighter standards of transparency, reproducibility, and accessibility (15–18). Agent-based models in particular offer advantages arising from their ability to capture correlated heterogeneities, for example between socio-economic status, geographical location, and access to healthcare.

In this paper, we introduce software for creating, calibrating, and analyzing agent-based models of HPV and cervical disease, called the Human papillomavirus simulator (HPVsim). HPVsim is built upon the architecture and design of the Starsim modeling platform, a flexible Python-based framework for epidemiological modeling which thus far includes Covasim for modeling COVID-19 (19), FPsim for family planning (20), and Synthpops for population modeling (21). Because this core software architecture has been described in detail elsewhere, in this paper we mainly focus on the two key innovations of HPVsim: (1) on the science side, HPVsim is designed to complement the existing suite of HPV models by offering a novel methodological approach to modeling HPV’s natural history; and (2) on the software side, we introduce a multi-scale modeling technique that substantially decreases the variability of agent-based simulations. To test the scientific realism of HPVsim, we created a model of HPV transmission in India in collaboration with the National Disease Modeling Consortium of India. The details of this model are interwoven throughout the following section on design and implementation, and additional examples of HPVsim’s use are contained in the results section along with validation of the multi-scale approach.

## 2. Design and implementation

### 2.1 Populations and networks

#### 2.1.1 Overview

HPVsim models agents (i.e., people) over time, simulating the transmission of HPV within sexual networks and the progression of HPV to cervical dysplasia and cervical cancer within individuals. A simulation is initialized with a population of agents, typically using HPVsim’s ‘location’ argument to select a distribution of ages and sexes for a particular country (19). Demographic updates are made annually to account for births, deaths, and migration (Table S1). HPV is transmitted between sexually active agents over sexual networks, which can be configured by the user by adjusting various input parameters (Table S2). The sexual networks are generated by an algorithm described in detail in Table S3, but which can be briefly summarized as a female-driven partner selection algorithm that assigns males to female partners based on age and other preferences (e.g., type of relationship, concurrency, and geospatial location), and then determines the relationship duration, propensity for condom use and frequency of sex for each relationship based on the age and preferences of both parties. Similar kinds of agent-preference algorithms have been used in other agent-based models (23), and we chose to follow this approach here because it allows the network to be more easily parameterized with respect to available sexual behavior data (as compared to using a mechanistic topology, for example).

#### 2.1.2 Example: parameterizing a sexual network for India

To create a sexual network model for India, we used data from the Demographic and Health Surveys (DHS) Program to inform the distribution of the ages of sexual debut for both males and females (Figure S1A), and adjusted the marriage incidence rates (parameter ‘layer_prob’ in the model) to match reported DHS data on the proportion of women married by age (Figure S1B). According to the DHS, very few women in India have extra-marital partners. To capture this, we used a negative binomial distribution for the degree distribution of female’s casual contacts, with parameter values chosen so that ∼90% of women have no non-marital partners (Figure S1D).

### 2.2 HPV transmission

HPVsim models HPV transmission within serodiscordant partnerships, with per-act probability *β*_*g*_ for genotype *g*. By default, HPVsim captures three genotype groups: types 16, 18, and a pooled group of all other oncogenic types. This latter group can also be disaggregated into the five types preventable by the 9-valent vaccine (“Hi5”, consisting of 31, 33, 45, 52, 58) and other high-risk types (OHR, consisting of 35, 39, 51, 56, 59), and we use these two genotype groups along with types 16 and 18 for the examples shown throughout this paper. For simplicity, we assume that individuals can only have a single infection with a given genotype at a time, i.e. a person currently infected with genotype *g* cannot get a second infection with the same genotype. We do, however, allow multiple concurrent infections with different genotypes. In the absence of clear data to inform within-host genotype competition and given the delay in seroconversion and antibody development to provide own- and cross-immune protection (24,25), we assume HPV infections are independent; agents are no more or less likely to acquire an infection of a different genotype if they are currently infected. The probability of a person *i* infected with genotype *g* transmitting to a susceptible person *j* within a given time step can be written as:

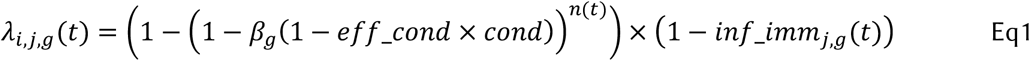

where *eff_cond* is the assumed efficacy of condoms (set to 50% by default), *cond* is the probability of condom use within this partnership, and *n(t)* is the number of sexual acts within this partnership between time *t* and time *t+dt*. The parameter *inf_imm*_*j,g*_*(t)* describes the person *j*’s protective immunity to infection against genotype *g*, which can be conferred by either infection (Table S4) or vaccination (Table S5). We assume men do not acquire neutralizing immune protection after clearing infection, based upon a meta-analysis of natural acquired immunity (26).

### 2.3 Disease natural history

#### 2.3.1 Motivation and overview

Historically, cervical disease was classified into histological grades depending on how much of the overall thickness of the epithelium contained neoplastic cells (CIN1 = basal third, CIN2 = up to two-thirds, CIN3 = more than two-thirds). This classification system meant that data from HPV screening programs could be used to estimate transition probabilities between these states, and these transition probabilities form the basis of the most frequently used set of existing published HPV models (27).

When designing a natural history model for HPVsim, our primary aim was to decouple the model’s representation of disease progression from the particular type of screening method or clinical classification system used, since both of these have evolved substantially over the past decade (e.g., the above-mentioned histological grades are no longer recommended) and continue to evolve as new methods are made available e.g., AI-based visual evaluation tools (28,29). Therefore, in HPVsim we model a continuous process of disease progression. In brief, the algorithm steps are as follows. First, we sample the duration of HPV infection prior to either clearance or the onset of high-grade squamous intraepithelial lesions (HSILs; Figure 1A). Longer infections are more likely to result in the development of HSIL (Figure 1B). We then sample the duration of HSIL prior to clearance or cancer (Figure 1C). The longer that a woman sustains HSIL, the more likely it is that she will develop cancer; this is reflected in Figure 1D, which should be interpreted as the probability that a woman with HSIL that persists for more than X years developing cancer, noting that this does not specify at what point the cancer will commence since this is dictated by the durations given in Figure 1C.

**Figure 1.**
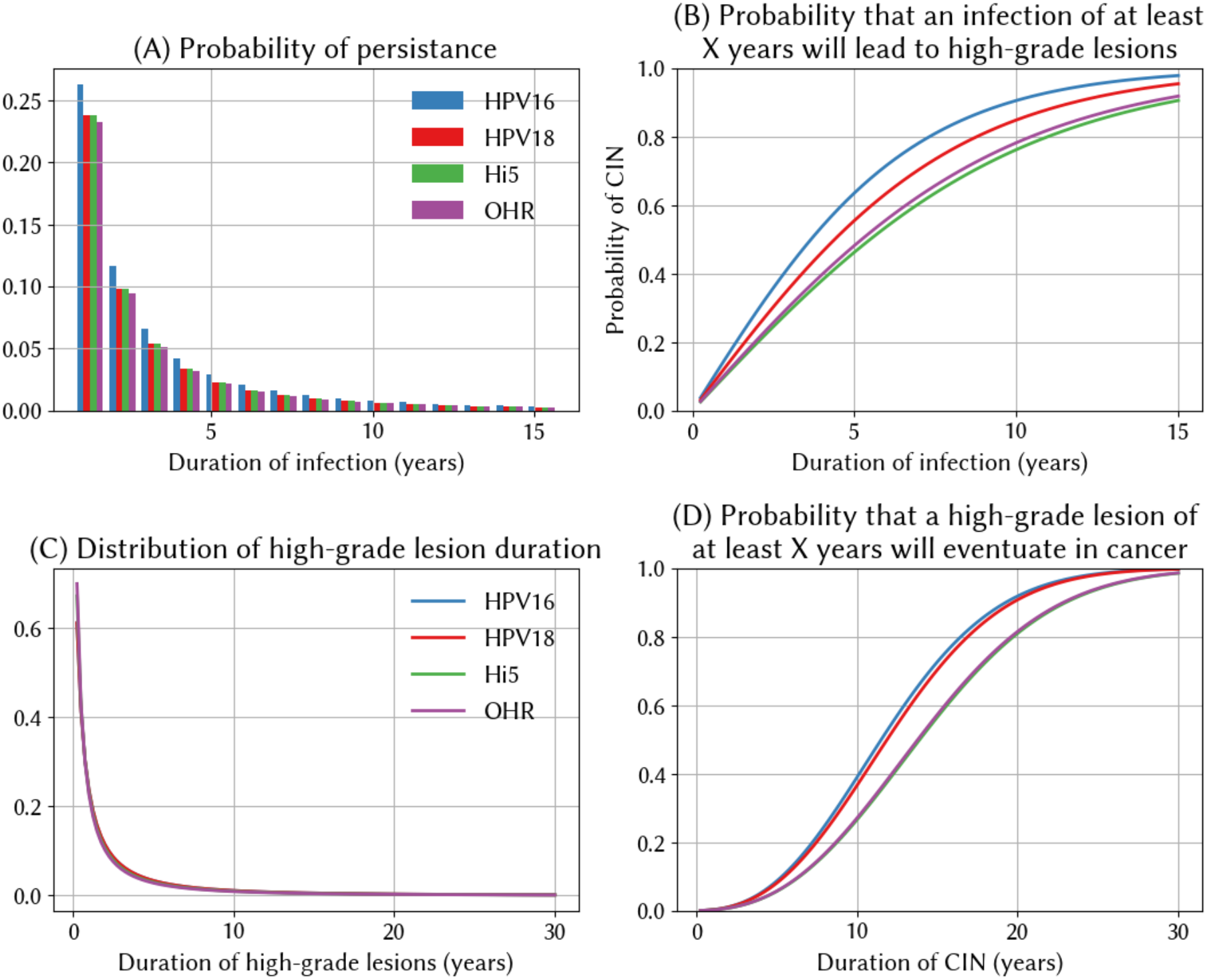
Infection and progression dynamics for types 16, 18, HI5 (a pooled group consisting of types 31, 33, 45, 52, and 58), and OHR (a pooled group consisting of 35, 39, 51, 56, and 59). (A) The duration of HPV infection prior to either clearance or the onset of high-grade squamous intraepithelial lesions (HSIL) follows a log-normal distribution, with mean values varying by genotype. (B) Mean relationship between the duration of infection, genotype, and the probability of developing HSIL. (C) The duration of HSIL prior to either clearance or the onset of cancer follows a log-normal distribution, with mean values varying by genotype. (D) Probability that HSILs lasting for at least X years will lead to cancer at the end of the duration drawn from the distribution in panel C.

#### 2.3.2 Choice of functions

For all three components of our natural history model – the distribution of infection duration, the duration-to-severity function, and the probability of cancer beginning – we need a functional form and default parameters. In this section we describe the functional forms (same for all genotypes), while the next section describes the parameter values (which vary by genotype).

For the distribution of the duration of infection, we use a log-normal distribution, since parameterizing this with respect to available data is straightforward (see next section). For the duration-to-HSIL function, we first consider the properties that this function should have. Available evidence suggests that women tend to spend relatively less time with no or clinically insignificant lesions, and longer with higher-grade lesions (30–32). This can be parsimoniously modeled using the concave part of a logistic function, such that the probability of an HSIL with genotype g after t years in individual *i* is:

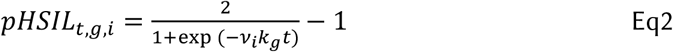

where *ν*_i_ is an individual effect that accounts for the fact that not all infections will progress at the same pace, and is drawn from a normal distribution with mean of 1.

We again use a log-normal distribution for the distribution of the duration of HSIL, with parameterization described in the next section. Finally, we need to specify a model of the probability of HSIL progressing to invasive cervical cancer. This is a binary outcome and depends on the extent and persistence of dysplasia in the cervical cells, so we use a standard geometric distribution:

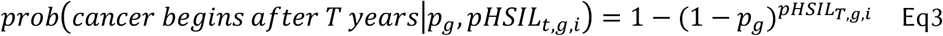

The parameter *p*_*g*_ represents the probability of a given cell becoming cancerous at any point in time, while the exponent *pHSIL*_*T,g,i*_ represents the cumulative probability of dysplasia (i.e., the integral of Equation 2). The exact form of *pHSIL*_*T,g,i*_ is given by:

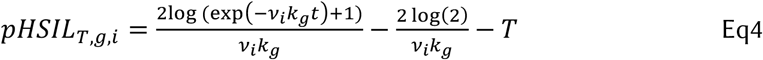

The function in Equation 3, while complex at first glance, has three features to recommend it. Firstly, because it’s a smooth approximation to a ramp function, it implies that there is an almost-zero probability of cancer beginning within the first few years of an infection, in line with the consensus understanding of the timeline to the onset of cervical cancer (30–32). Secondly, it has an intuitive interpretation as the cumulative HSIL-time of an infection (i.e., the integral of Equation 2). And finally, it does not rely on any additional parameters aside from those already used in Equation 2.

#### 2.3.3 Example: parameterizing the natural history model for India

All in all, our method for modeling HPV’s natural history has 6 parameters per genotype: the mean and standard deviation of the duration of pre-HSIL infection, the growth rate k_g_ in the infection-to-HSIL function, the mean and standard deviation of the duration of HSIL, and the per-cell probability of becoming cancerous at any given point in time.

For the distribution of HPV 16 infection durations, we use default values which imply that ∼50% of infections clear within 12 months and 60% within 24 months, in line with the findings from the Guanacaste cohort study (32). Values for remaining genotypes are listed in Table S6.

To find values for k_g_, p_g_, and the mean and standard deviation of the duration of HSIL for India, we used the sexual network described in Section 2.1.2 and estimated the natural history parameters by calibrating to cervical cancer cases by age, the distribution of HPV types found within women with precancerous lesions and the distribution of HPV types invasive cervical cancer. All data was taken from Globocan 2020 estimates and HPV Information Centre compiled meta-analyses. We ran 10,000 trials searching for parameters that minimized the sum of the normalized absolute differences between the model and these data. This resulted in parameter values documented in Table S6, with model fit shown in Figure S2, and comparison to Globocan estimates in Figure S3.

Using these estimate parameter values, we obtain the natural history model shown in Figure 1, including the severity function in Figure 1B, the probability of cancer shown in Figure 1C. Figure 2 then shows some additional validation plots for our natural history model of India. The dwell times in each state shown in Figure 2B, and are in line with other models (27). The share of women who would be observed in a given health state X years after their initial infection is plotted in Figures 2C and 2D, and is in line with available observational data (31). In separate work, we further validated our model by applying these estimates to 30 additional countries (33), under the expectation that the natural history should not vary substantially across settings.

**Figure 2.**
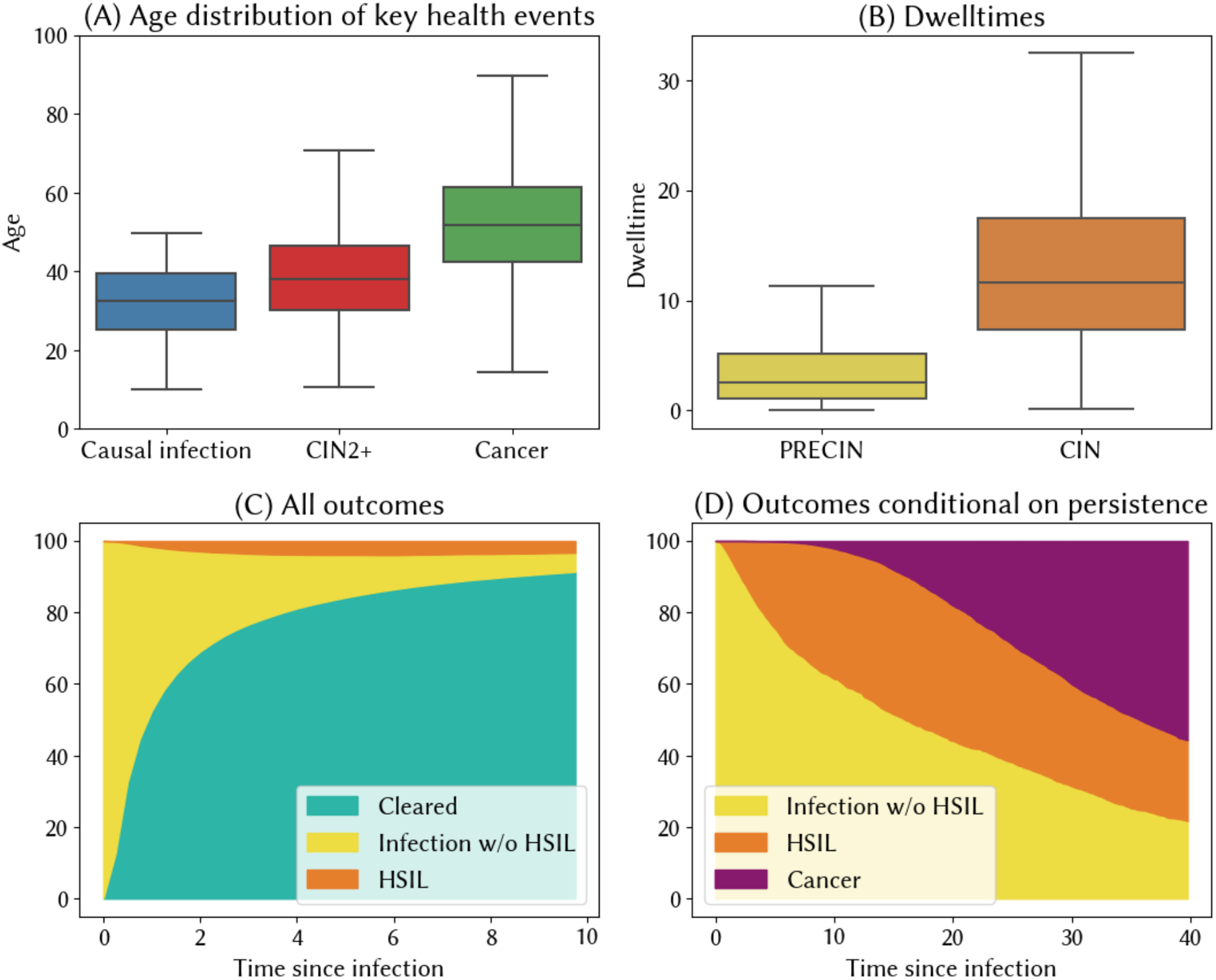
Implied outcomes from HPVsim’s natural history model for India. (A) Age distribution of key health events in the lead-up to cervical cancer, (B) Dwelltimes for no or low-grade lesions and high-grade lesions, for women who progress to cancer. (C+D) Estimated distribution of outcomes for all HPV infections (C), and conditional on persistence (D).

#### 2.3.4 Other implementation details

##### Performance

To optimize performance, we determine all prognoses upon infection, including the precise time points on which a woman will either clear infection or eventually progress to invasive cervical cancer. These prognoses can be updated at later dates in response to interventions (e.g. screening and treatment) or changes in a state of immune compromise, such as untreated HIV infection. Pre-specifying prognoses in this way means that we only need to change the health state of a small subset of women in each timestep.

##### Men

For men, a duration of infection is sampled from a log-normal distribution and date of clearance assigned at the time of infection. We do not model persistent infections in males or any associated precancer/cancer outcomes.

##### Immunity

There is some evidence to suggest that women who clear an HPV infection and are reinfected have a reduced probability of persistent infection or high-grade dysplasia (34). We capture this using an immunity parameter that reduces the average duration of re-infections of the same type, and also confers some cross-immunity (Table S4).

##### Age

There is some debate about the extent to which age plays a role in exacerbating the likelihood of an infection persisting or progressing to high-grade lesions (35). In HPVsim, we do not include an age-modifying effect by default, but we do include the option to add one if desired.

##### HIV

HPVsim can also capture the impact of HIV infection on HPV acquisition and natural history. At each time step, individuals in the simulation can be exogenously infected with HIV based upon their age, sex and the year of the simulation. Upon infection, the ART adherence of that agent is determined, which will then be used to modify any future risk of HPV acquisition and progression. While individuals adherent to ART can fail to suppress HIV or experience immune reconstitution, for simplicity we assume an individual’s immune state is directly proportional to adherence. At the point of HIV infection, for individuals with prevalent HPV infection, the prognoses are modified using relative risk estimates derived from a meta-analysis (Table S9).

### 2.4 Interventions

The central pillars of the global public health response to HPV consist of prophylactic vaccination programs, cervical screening programs, and treatment of both precancerous lesions and cancer. HPVsim allows users to model these standard interventions (Table 1), and additionally to define their own custom ones that might reflect country-specific strategies for prevention and treatment. The general design of the standard interventions (screening, triage, treatment, and prophylactic vaccination) is intended to allow users to flexibly specify both the product and the details of its delivery.

**Table 1.** Overview of interventions included in HPVsim.

| Intervention | Core assumptions, all user-modifiable |
| --- | --- |
| Prophylactic vaccination | **Efficacy:** a single dose of a prophylactic vaccine is assumed to confer neutralizing immunity to the genotypes that it is specifically targeted at, with efficacy of 97.5% (36), as well as lower levels of cross-immunity to other genotypes (Table S5)<br>**Duration of protection:** By default, no waning of immunity (37–39), with an option to include waning.<br>**Delivery:** Flexibly defined by the user; see docs examples. |
| Screening and triage | **Screening products:** HPV DNA testing, cytology, and visual inspection with acetic acid (VIA) have been built into HPVsim, using sensitivity and specificity assumptions documented in Table S7. Other products can be added following examples provided in the model documentation.<br>**Delivery:** Flexibly defined by the user; see docs examples. |
| Treatments for precancer | **Treatment products:** Thermal ablation is assumed to be 93% effective at removing lesions (40,41); excisional treatments are assumed to be 95% effective at removing lesions (42); both are assumed to be 80% effective at clearing infection (43,44).<br>**Delivery:** Flexibly defined by the user; see docs. |
| Treatments for cancer | Radiation therapy is included with HPVsim as a possible treatment option, which acts to extend the lifespan of people with invasive cancer. Chemotherapy and surgery are not included by default, but can be added as custom interventions. |
| Therapeutic vaccination | **Therapeutic vaccine products:** There are currently no |
|  | therapeutic vaccine products on the market, although at least one clinical trial is underway (45,46). HPVsim includes basic infrastructure for modeling therapeutic vaccination. The default assumptions are that the therapeutic vaccine will be delivered as a two-dose regimen targeted at HPV16 and 18, with the two doses in combination having 50% efficacy at regressing high-grade lesions and 90% efficacy at virological clearance. Different assumptions can be explored by changing these parameters.<br>**Delivery:** Flexibly defined by the user; see docs for examples of mass vaccination and vaccination within screening programs. |

### 2.5 Multiscale modeling

In agent-based modeling, it is common for one “agent” to represent more than one person (or more precisely, for agents to represent a statistical sample of the population, much like in a survey). However, a problem arises when some events being simulated are very frequent (such as HPV infection, affecting a majority of people), and other events are very rare (such as developing cervical cancer, affecting roughly 1 in 150 women). Under these circumstances, agent-based modeling can produce considerable stochastic variation, especially with smaller simulated population sizes. To illustrate this, consider using a binomial distribution to estimate the expected number of cases in a given year. With a population size of 10k agents, our best estimate would be 66 cases, with a 95% prediction interval of 48-86 cases. From year to year and simulation to simulation, we would observe a high degree of variability, with a coefficient of variation (CoV) of 0.12. Under this simplified framework, a tenfold decrease in the simulated population size translates to a √10-fold increase in the coefficient of variation in the estimated number of cases. Avoiding large variability would therefore typically require large population sizes. To avoid this, HPVsim uses a novel approach called *multiscale modelling* to optimize computational efficiency.

In a multiscale modeling approach, different types of agents are modeled with different levels of “resolution” or “sampling.” For example, for a population of 10 million people (equivalent to a medium-sized country) being modeled, specifying 100,000 agents is roughly the number of agents required to provide a good tradeoff between statistical accuracy and simulation speed. This means there is a scale factor (or “weight”) of 100 for each agent (or put another way, people are downsampled by 100). We call these “Level 0” agents since they are at the lowest (default) level of resolution. These are the agents who participate in the sexual mixing network, who are able to contract and transmit HPV, and who can give birth to new agents. “Level 1” agents have a lower scale factor (higher resolution, lower weight per agent), and correspond to agents who will (without intervention) develop cancer. These Level 1 agents all have HPV (we adjust to prevent double-counting), but do *not* participate in the disease transmission network; instead, only their disease progression is tracked.

When an agent becomes infected with HPV, their disease duration and outcomes are pre-calculated (although these can be later modified through screening, treatment, and other interventions). This is when multiscale modeling is applied. A schematic illustration of the algorithm is shown in Figure 3, with the complete algorithm outlined in Table S8.

**Figure 3.**
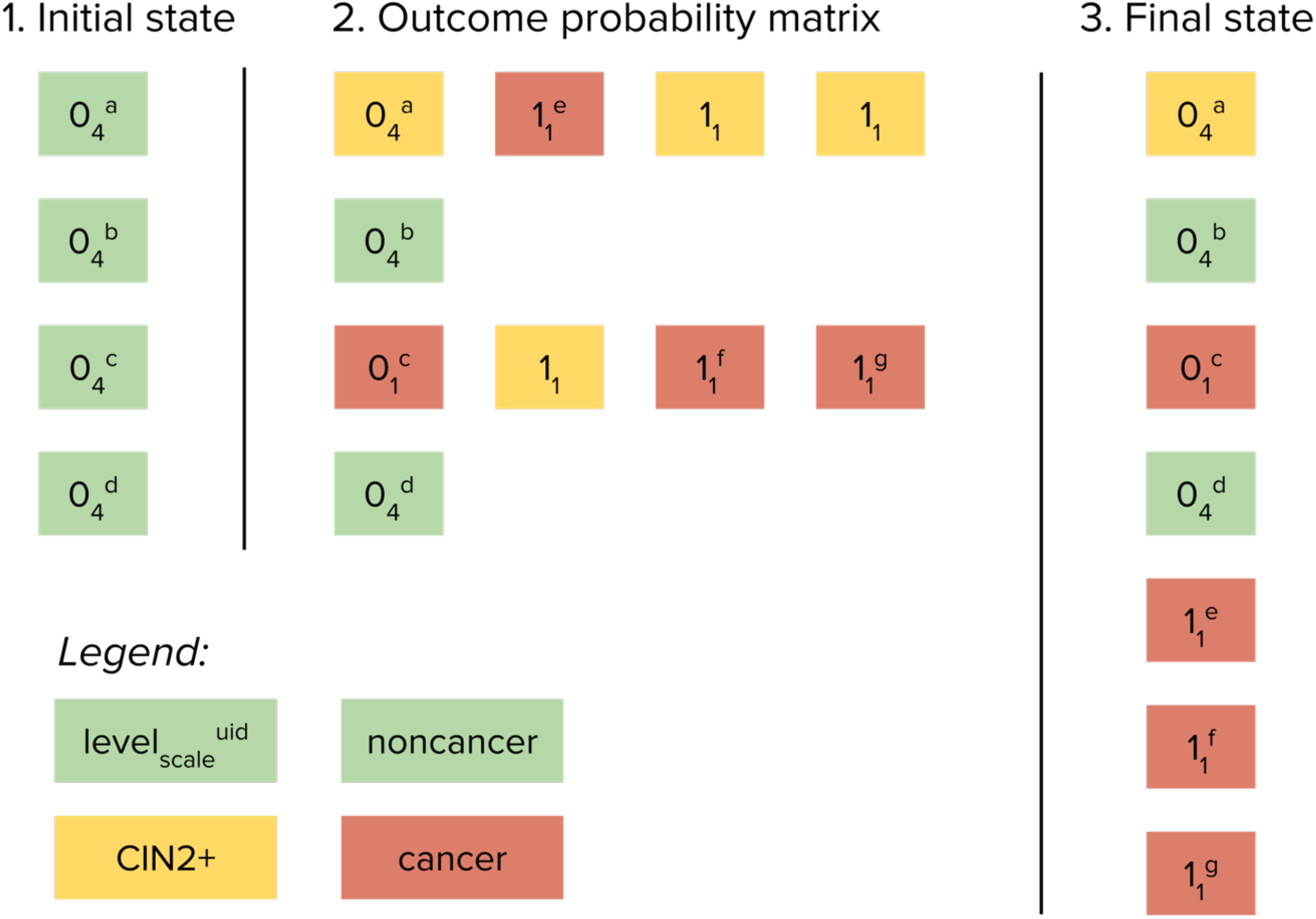
Schematic example of multiscale modeling implementation. In this illustration, there are *N*=4 agents, the probability of developing CIN2+ is 50%, the probability of developing cancer given CIN2+ is 50%, the scale factor (“weight”) for level 0 agents (non-cancer, green) is *S*_*0*_*=*4, the scale factor for level 1 agents (cancer, red) is *S*_*1*_*=*1, and therefore the multiscale agent ratio is *S*_*0/*_*S*_*1*_*=*4. On the initial step of the algorithm, there are four agents, each with a scale factor of 4, representing 16 people without cancer. On the second step, two level 0 agents (with UIDs [universal identifiers] *a* and *c*) develop CIN2+; one of these agents (*c*) then develops cancer, and has her scale factor changed from *S*_*0*_*=*4 to *S*_*1*_*=*1. Both CIN2+ agents then generate three new *potential* level 1 agents; of these six new potential cancer agents, three develop cancer and become level 1 agents (with UIDs *e, f*, and *g* assigned at the point of creation). On the third step, the four original level 0 agents (including one with CIN2+ and one with cancer) are combined with the three new level 1 agents (all with cancer) to form the new list of *N*=7 agents. Note that although there were *N*=4 agents on step 1 and *N*=7 agents on step 3, the sum of the scale factors (weights) of the agents remains the same, i.e. 16.

## 3. Results

### 3.1 Variance reduction from multiscale modeling

In Figure 4, we compare two key simulation features – variability and simulation time – as a function of the multiscale agent ratio and the number of agents in the simulation. To produce this figure, we ran ten simulations for each combination of the multiscale agent ratio and the number of agents, and computed the coefficient of variability (CoV) in deaths and the overall average time per simulation. As an example, suppose we were running a simulation with 50k agents without multiscale (the second largest dark blue dot on Figure 4). This would take ∼15 seconds to run and produce a CoV of 0.028 across 10 simulations. To decrease variability, we could either increase the number of agents or increase the multiscale ratio. If we increased the number of agents to 100k, our simulation would take almost twice as long (∼28 seconds) and would produce a CoV of 0.023 (the largest dark blue dot on Figure 4). By contrast, if we increased the multiscale agent ratio to 100, we would see a slightly smaller increase in simulation time (to 26 seconds), but the CoV would decrease by much more, to 0.007 (the second largest yellow dot on Figure 4).

**Figure 4.**
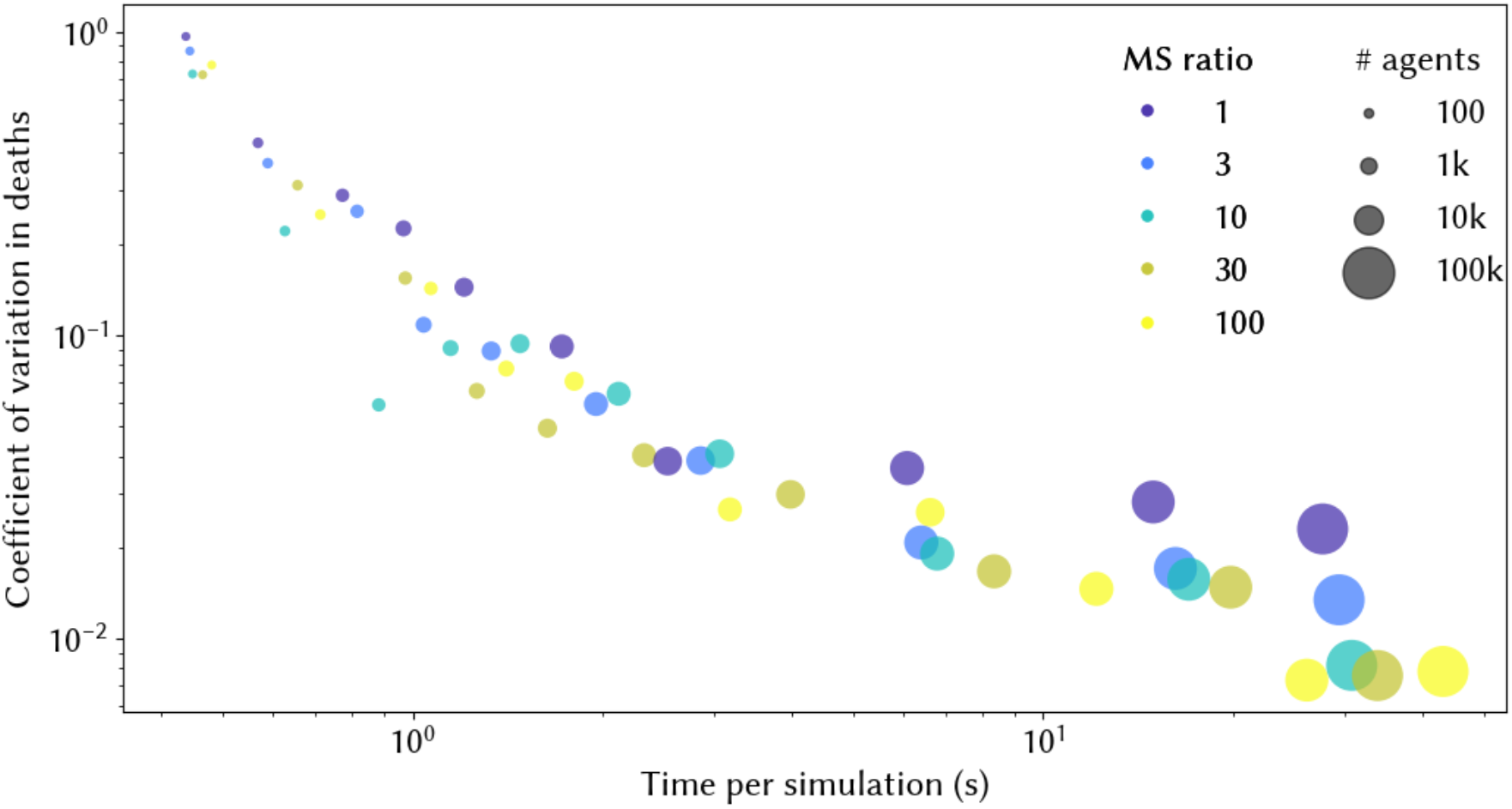
Trade-off between variability (measured by the coefficient of variation in deaths) and simulation time. Simulation times are calculated as the average across 10 simulations. MS ratio = multiscale ratio, and represents the ratio between the number of people represented by agents without cancer and the number represented by agents with cancer (see Figure 3).

### 3.2 Representing diverse sexual networks

For this example, we created two different sexual networks to highlight how patterns of sexual behaviour affect HPV epidemiology and outcomes. The first represents a setting where premarital sex is relatively common, which we model by assuming a Poisson distribution for women’s preferred number of partners, with 80% of women preferring to have at least one casual partner. The second represents a setting where premarital sex is very rare, and 90% of women have no non-marital partners, i.e. similar to the model for India that we developed throughout Section 2. In order for HPV to transmit in this second setting, we assume significant overdispersion in the degree distribution for females, i.e. a small number of females have a higher number of partners. We capture this using a negative binomial distribution for women’s preferred number of partners, parameterized with n=0.025 and probability of success p=0.0125. In each setting, we will assume that 25% of married males and 2.5% of married females have extra-marital relationships, and in each setting we assume the age of sexual debut is log-normally distributed with a mean of 17 for females and 20 for males, and a standard deviation of 2 for both sexes.

In Figure 5, we plot the degree distribution for each setting, and alongside this we plot the distribution of ages at which key health events in the lead-up to invasive cancer take place. In the setting where premarital sex is common, the median age at which women who eventually proceed to invasive cervical cancer acquire their casual HPV infection is 25 years. However, it is 5 years older in the setting where premarital sex is rare. Without widespread premarital sex, there are a small share of sexually active women who acquire infection at a young age, but the majority of women acquire infection later, after their spouse transmits an infection acquired from an extra-marital relationship. This would have implications for the ideal age to target for cervical screening programs, as well as delivery of other interventions like a therapeutic vaccine.

**Figure 5.**
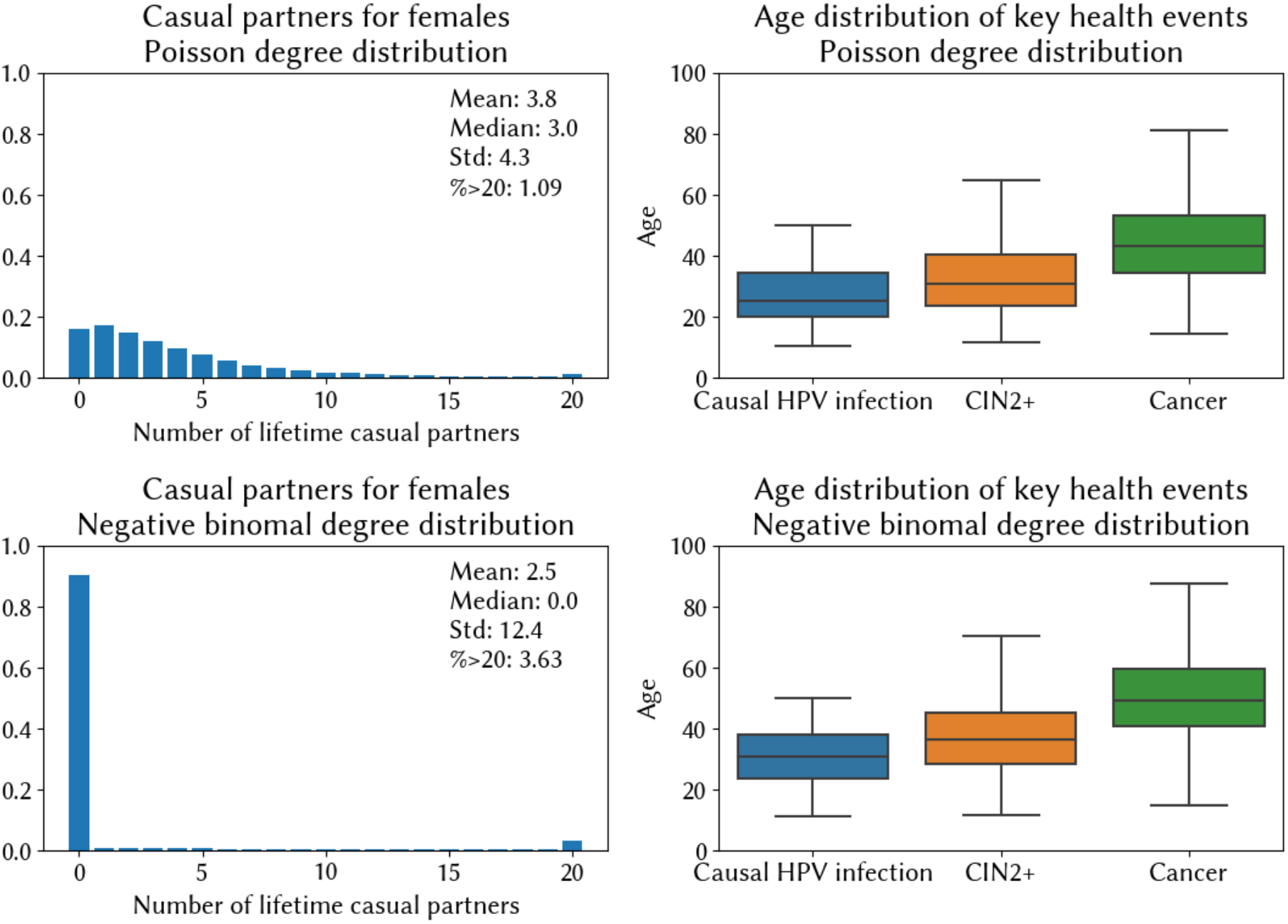
Relationship between the sexual network and the age of causal HPV infection. The top row describes a setting where pre/extramarital sex is common, while the bottom row describes one where very few women have sexual partners outside of marriage.

### 3.3 State-level variability in cervical cancer incidence and screening in India

There are large differences in reported age-standardized rates of cervical cancer incidence across the states of India, ranging from 4.8 cases per 100,000 women in 2020 in Assam, up to 27.7 cases per 100,000 women in Papumpare (47). These differences cannot be attributed to HPV vaccination programs, since these only commenced in 2018 and have thus far been limited to two districts of Punjab, opportunistic vaccination in Delhi, and one state-wide program in Sikkim (48). In September of 2023, we convened a training workshop at the Indian Institute of Technology, Bombay, where members of the National Disease Modeling Consortium speculated on possible causes for the state-level variation. Among the many possible contributing factors (including wide variations in demographic, behavioral and reproductive risk factors), one additional factor raised as a possible explanation for this phenomenon was variation in screening uptake. Although all states report relatively low lifetime screening coverage (47), these data are based on usage of the public system, and in some states utilization of the private health system may be higher, meaning that screening rates could be higher than observed in the data.

For this illustrative case study, we investigated how much cervical cancer incidence would vary as a function of screening coverage levels alone, holding all else equal. We simulate a population with the demographics and sexual behavior of India, as presented throughout Section 2. We then assume that screening would have been available as an out-of-pocket service starting from the year 2000, with uptake scaling up linearly from 0% to reach 5, 10, 15, or 20% over the subsequent 20 years. For each scenario, we run 20 simulations and take the average and the 90% range to account for stochastic differences. We find that prolonged screening programs, even with relatively low levels of coverage, may result in lower levels of age-standardized cancer incidence. We estimate an ASR of cancer incidence without screening of 17.17 [90% range: 15.86, 19.45] in 2020; if we assume 20% lifetime screening coverage by 2020 this falls to 12.39 [90% range: 11.73, 14.03] (Figure 6). Given that 20% would represent extremely high uptake of the private system, we conclude that while some of the state-level differences may be attributable to private sector screening, it is unlikely to be the main driver of state-level variation.

**Figure 6.**
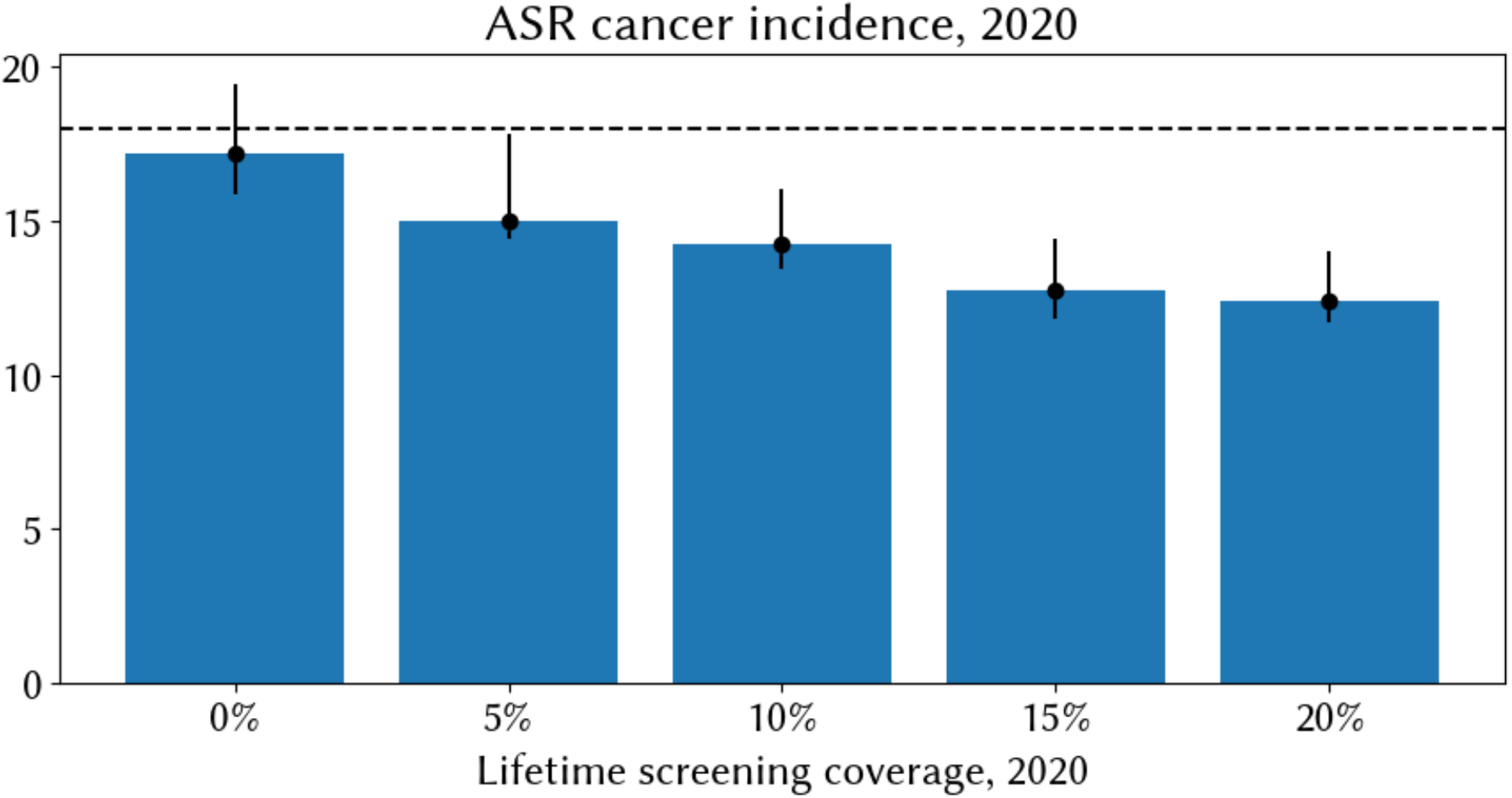
Median modeled age-standardized rate of cervical cancer incidence in 2020 across 20 simulations (blue bars) and 90% range (black bars). Modeled results are based on a setting representative of India but with varying hypothetical levels of historical screening coverage, increasing linearly from 2000 to reach the values displayed on the x axis in 2020.

## 4. Availability and future directions

### 4.1 Availability

The source code for HPVsim, including documentation and tutorials, is freely available via <u>GitHub</u>. The repository containing the India model calibration is <u>here</u>. The scripts to produce the figures and analyses used in this paper are in a separate Github repository available <u>here</u>. This manuscript refers to all features and parameters of version 2.0.0 of HPVsim. HPVsim follows best practices for version and dependency management, including a CI/CD/CT pipeline with comprehensive code coverage to ensure that no regressions are introduced as dependencies are updated.

### 4.2 Future directions

The HPVsim software can be used to rapidly produce models of HPV and cervical cancer, but before a model can be used within a given setting, it must first be calibrated, i.e. contextualized by identifying parameters that generate model outputs resembling observed data. The calibration process represents an important step for future users and developers of HPVsim. Within this paper we presented a use case demonstrating how this can be done; further examples can be found in related studies (21,22), and example scripts for setting up and running a calibration can be found in the Documentation and Tutorials. However, an important future direction for HPVsim research is to demonstrate its ability to fit to more diverse data sets, and to evaluate post-hoc out-of-sample fit. In addition, the model parameters could be further improved with access to better HPV burden and cervical cancer mortality estimates over time and by region.

The sexual network is an important area for future development. First, HPVsim currently only models heterosexual partnerships and cisgender individuals; expanding this could potentially increase the accuracy of model outputs. Second, implementing assortative mixing by geographical location or demographic factors other than age groups would help understand the impact of network topology on disease transmission. Furthermore, heterogeneities in individuals’ risk of cervical disease and/or uptake of interventions may also correlate with the assortative mixing pattern in sexual networks. Incorporating functionalities to customize and evaluate such heterogeneities and their correlation with network structure would be key for identifying core groups for intervention.

The calibrated model for India that was presented throughout this paper is being further developed by members of the National Disease Modeling Consortium of India, with a particular focus on deeper investigation of the state-level variation in cervical cancer incidence, and how this variation may influence optimal vaccination policies for each state. There have been some studies on the cost effectiveness of vaccination and screening in India (49), but none have captured the variation by state, a detail that is likely to be vital for motivating programmatic implementation.

While HPVsim’s structure is generic enough that theoretically the model can be applied to any country, we have designed it with lower- and middle-income countries in mind, and as such it may not capture many of the downstream features specific to higher-income countries (e.g. we do not yet model cancer progression or treatment in detail). HPVsim’s sexual network currently only includes partnerships between men and women. We do not capture diseases caused by HPV apart from cervical cancer (e.g. genital warts, and cancers of the vulva, vagina, penis, anus, and oropharynx), although these could be integrated into future versions by adapting the existing framework. Although we do model heterogeneities in individuals’ risk of cervical disease, we do not specifically link these varying risk levels to known confounders such as parity, tobacco use, and other behavioural factors. We also do not capture differences in health-seeking behaviour and uptake of interventions arising from age, race, ethnicity, gender identity, sexuality, or income. All of these current limitations of HPVsim represent possible avenues for future development contributions.

### 4.3 Concluding statement

In response to the continued evolution of policy recommendations around HPV vaccination, screening, and treatment, we have created a model which, over time and as it is further applied, will enable researchers and policy makers to analyze new technologies, evaluate different strategies, and feed into decision-making pipelines. Models like HPVsim are often used to evaluate the impact of interventions, information which can then be incorporated into a cost-effectiveness analysis if cost data are available. We are committed to ongoing development and improvement of HPVsim in partnership with stakeholders, collaborators, and users of the model, not only because such collaborations strengthen the model itself, but more importantly, because they provide an opportunity to work together towards the fundamental goal of cervical cancer elimination.

## Data Availability

The source code for HPVsim, including documentation and tutorials, is freely available via GitHub (hpvsim.org). The repository containing the India model calibration is available at https://github.com/hpvsim/hpvsim_india. The scripts to produce the figures and analyses used in this paper are in a separate Github repository available at https://github.com/hpvsim/hpvsim_methods_manuscript. This manuscript refers to all features and parameters of version 2.0.0 of HPVsim. HPVsim follows best practices for version and dependency management, including a CI/CD/CT pipeline with comprehensive code coverage to ensure that no regressions are introduced as dependencies are updated.

## Acknowledgments and contributions

### Author contributions

Conceptualization, software, methodology, and investigation: RMS, JAC, CCK, and RGA. Formal analysis: MZ. Data curation: MCB. Validation: SL, LY. Supervision: DJK, DWR. Writing – original draft: RMS, JAC, CCK. Writing – review & editing: all authors.

## Acknowledgements

HPVsim model development has been led by Jamie Cohen, Robyn Stuart, Romesh Abeysuriya, and Cliff Kerr. This project is sponsored by Hao Hu and Daniel Klein. We thank our expert advisers for their frequent and helpful consultations on model conceptualization and development. These include but are not limited to Sharon Achilles, Maike Scharp, Celina Schocken, Peter Dull, John Schiller, Chris Karp, and Holger Kanzler. Mark Jit, David Wilson, Sherrie Kelly, Jasmina Panovska-Griffiths, Andrew Shattock, Edinah Mudimu, Michelle O’Brien, Helen Olsen, and Greer Fowler provided editing and informal review. Erin Ingle provided valuable literature review support to inform model parameters. Tremendous gratitude to Mandy Izzo for her graphical content and editing support.

## Additional information

For more information or to get involved, please.

