## Supplementary materials for "HPVsim: An agent-based model of HPV transmission and cervical cancer"


| | |
|---|---|
| Table S1: Demographic inputs | 2 |
| Table S2: HPVsim parameters and default values | 3 |
| Table S3: HPVsim sexual network | 9 |
| Figure S1: Sexual network for India | 10 |
| Table S4: Cross-immunity matrix following infection | 11 |
| Table S5: Prophylactic vaccine parameters | 12 |
| Table S6: HPVsim genotype parameters and default values | 13 |
| Figure S2: Fitting the model to data from India | 14 |
| Table S7: Sensitivity/specificity of screening products | 15 |
| Table S8: Multi-scale modeling algorithm | 16 |
| Table S9: HIV parameters | 17 |
| References | 18 |

### Table S1: Demographic inputs

| Data type | Notes | Source |
|---|---|---|
| Births | New births are added to the simulated population using crude birth rates by year as published by the World Bank. | (1) |
| Deaths | Age specific mortality rates by sex and year are applied to the simulated population using rates included with the UN's 2022 World Population Prospects (WPP). | (2) |
| Migration | After adding births and removing deaths, we compare the overall modeled population size to the projected population size, which is taken from the UN WPP's mid-fertility projections. Discrepancies are assumed to be attributable to migration, so the final update consists of either adding or removing people from the simulation. Both methods are applied to maintain historical and projected age-distribution over time. By default, when adding migrants to the population, we draw attributes such as age, immunity profile, and vaccination status from a distribution based on that of the existing population, although these attributes can be changed if desired. | (2) |

Table S1 notes:

1. All data required for this workflow are automatically pulled from World Bank and UN WPP sources cited above upon installation of HPVsim, using data scrapers that generate and locally store object files with demographic data for all countries in the world. When new data are released, these will be included in future releases of HPVsim, and will automatically update for users when they update their HPVsim version.

### Table S2: HPVsim parameters and default values

| Parameter | Code name | Description / sources | Default value |
|---|---|---|---|
| **Population parameters** | | | |
| Number of agents | n_agents | Number of agents at the beginning of the simulation. With population growth, this number increases over time | 20,000 |
| Total population | total_pop | The size of the real world population that the simulated population is assumed to represent | If a location is provided, this is automatically populated with the population size of the specified location at the beginning of the simulation. If not, it is assumed to be the same as n_agents unless a different value has been given |
| Population scale factor | pop_scale | Defined as the ratio between total_pop and n_agents | Automatically calculated based on either the location (if provided) or total_pop. |
| Multiscale factor | ms_agent_ratio | Ratio of scale factor of cancer agents to normal agents (described in Section 2.4). | 1 by default |
| Sexual network structure | network | The two standard sexual networks are described in Table S3 | The 'default' network (Table S3) |
| Location | location | Geographical setting - determines the demographic inputs (Table S1) | None |
| Number of persons surviving to age x | lx | The number of persons surviving to exact age x (2) | Automatically populated based on location |
| Birth rates | birth_rates | Crude birth rate per 1000 (3) | Automatically populated based on location |
| Death rates | death_rates | Death rates | Automatically populated based on location |
| Relative birth rates | rel_birth | Convenience parameter for scaling the birth rates. | 1 (no scaling) |
| Relative death rates | rel_death | Convenience parameter for scaling the death rates. | 1 (no scaling) |
| **Initialization parameters** | | | |
| Initial HPV prevalence | init_hpv_prev | Distribution of HPV prevalence by age and sex in the beginning year | 18-24 years: 5% |

| | | | |
|---|---|---|---|
| | | of the simulation. The default values are all placeholder assumptions. | 25-33 years: 7%<br>34-43 years: 5%<br>44-63 years: 2%<br>65+ years: 1% |
| Initial HPV type distribution | init_hpv_dist | Distribution of HPV genotypes in the beginning year of the simulation | Equal distribution among simulated genotypes |
| Relative initial prevalence | rel_init_prev | Convenience parameter for scaling initial HPV prevalence values | 1 |
| **Simulation settings** | | | |
| Start year | start | Start year of the simulation | 1995 |
| End year | end | End year of the simulation | 2030 |
| Number of years | n_years | Number of years in the simulation | Calculated based on start and end years |
| Burn in period | burnin | Number of years to discard from the beginning of the simulation. Recommended 20-30 years to ensure that the sexual network has reached a stable state. | 25 |
| Timestep | dt | Simulation timestep for everything except demographics | 0.25 years (3 months) |
| Demographic timestep | dt_demog | Simulation timestep for demographics | 1 year |
| Random seed | rand_seed | Seed used for random number generation | 1 |
| Model verbosity | verbose | How much output to print when running the model. | 0.1 (meaning that output is printed every 10 timesteps) |
| Immunity waning assumption | use_waning | Whether or not to assume that immunity wanes over time. | False |
| Migration assumption | use_migration | Whether or not to approximate migration | True |
| HIV model setting | model_hiv | Whether or not to model HIV | False by default |
| **Network parameters** | | | |
| Sexual debut | debut | Default values are placeholder assumptions only, as this varies by setting. Typical data source: DHS. | Females: $N(18.5, 2^2)$, Males: $N(19.5, 2^2)$ |

| | | | |
|---|---|---|---|
| Relationship duration (years) | dur | Default values are placeholder assumptions only, as this varies by setting. Typical data source: DHS | Default network:<br>- Marital ($dur_m$): N(10, $2^2$)<br>- Casual ($dur_c$): N(2, $1^2$)<br>- One-off ($dur_o$): 0<br>Random network:<br>- All ($dur_a$): N(5, $3^2$) |
| Proportion with >1 partner of the same relationship type | partners | Default values are placeholder assumptions only, as this varies by setting. Typical data source: DHS | Default network:<br>- Marital ($conc_m$): 1%<br>- Casual ($conc_c$): 20%<br>- One-off ($conc_o$): 0%<br>Random network<br>- All ($conc_a$): 1% |
| Coital acts (per year) | acts | Within the model, coital frequency changes with age, and these values are assumed to be the number of acts for people at their sexual peak. Very little data is available, and these are assumptions. Users can change these distributions. As given here, the distributions are negative binomial distributions parameterized via two parameters representing the rate (mean) and dispersion. | Default network:<br>- Marital ($acts_m$): NegBin(80, 40)<br>- Casual ($acts_c$): NegBin(10 ,5)<br>- One-off ($acts_o$): NegBin(1, 0.1)<br>Random network:<br>- All ($acts_a$): NegBin(100, 50) |
| Condom usage | condoms | DHS data is typically available on condom usage at last act. | Default network:<br>- Marital ($cond_m$): 1%<br>- Casual ($cond_c$): 20%<br>- One-off ($cond_o$): 10%<br>Random network:<br>- All ($cond_a$): 25% |
| Participation rates | layer_probs | DHS data are available on the share of the population of each age group that are married. Participation in casual and one-off relationships are assumptions | Default network:<br>- Marital, casual, and one-off : varies by age<br>Random network:<br>- All: varies by age |
| Age mixing | mixing | DHS data are available on age differences within married couples. | Default network:<br>- Marital, casual, and one-off): varies by age<br>Random network:<br>- All: varies by age |

| | | | |
|---|---|---|---|
| Coital frequency by age | age_act_pars | The age of sexual peak is described by the *peak* parameter. The age of sexual 'retirement' is captured by the retirement parameter, and the *peak_ratio* and *retirement_ratio* describe how the number of acts that a couple at debut and retirement relates to the peak value.<br><br>For example, suppose a couple consists of a woman aged 40 and a man aged 50, so their average age is 45. We draw an annual number of acts for this pair from the distribution of *acts* (i.e., the parameter defined above, which represents the number at their sexual peak. With parameters peak=30, retirement=75, debut_ratio=0.5, and retirement_ratio=0.1, the actual number of acts that they are assumed to have at ages 40 and 50 is then calculated to be 60+(1-0.1)*60/(30-75)*(45-30)=42. The following year it will be 40.8, and so on until they reach ages 70 and 80, when it will stabilize at 6 acts/year. | Default network:<br>- Marital: peak=30, retirement=100, debut_ratio=0.5, retirement_ratio=0.1<br>- Casual and one-off: peak=25, retirement=100, debut_ratio=0.5, retirement_ratio=0.1<br>Random network:<br>- All: peak=30, retirement=100, debut_ratio=0.5, retirement_ratio=0.1 |
| **Basic transmission** | | | |
| Transmission probability | beta | Assumption; can be varied by genotype. | 0.05 |
| Relative transmissibility of insertive partners in penile-vaginal intercourse | transm2f | Default value taken from (4). | 3.69 |
| Efficacy of condoms | eff_condoms | Value taken from (5) | 0.7 |
| **Disease progression** | | | |
| Clinical cut-offs | clinical_cutoffs | Cellular dysplasia starts at the basal level and progresses outwards. The categorization of CIN1, CIN2, and CIN3 are based histologically on what proportion of the epithelial layer is dysplastic (the innermost 0-33% = CIN1; 33-66% = CIN2; and 66-100% = CIN3). These parameters reflect these clinical cut-offs. | CIN1: 33.3%, CIN2: 67.6%, CIN3: 99% |
| Probability of HPV infection progressing to latency | hpv_control_prob | There is ongoing scientific debate as to whether HPV clearance is really 'true' clearance, or whether it may represent latent control. This parameter captures the probability that a woman controls the infection latently. It is set to 0 by default but can be varied in sensitivity analyses. | 0 |
| Reactivation probability | hpv_reactivation | If a woman has controlled her HPV infection latently, this parameter | 0.025 |

| | | | |
|---|---|---|---|
| | | describes the annual probability of the infection reactivating. | |
| Duration of infection for males | dur_infection_male | Duration of infection for males | logN(1,1) |
| Severity distribution | sev_dist | Distribution to draw individual level severity scale factors | logN(1,0.1) |
| Duration of episomal infection before clearance or integration | dur_episomal | Varies by genotype | See Table S6 |
| Severity rate | sev_rate | Varies by genotype | See Table S6 |
| Transformation probability | transform_prob | Varies by genotype | See Table S6 |
| Duration of cancer | dur_cancer | Duration of untreated cancer. This distribution is chosen to roughly align with the 5-year survival rates published by Surveillance, Epidemiology, and End Results (SEER) (6). Estimates of cancer deaths in the absence of treatment will be lagged by a period determined by this parameter, and in practice this parameter may need to be adjusted during calibration to match data on the distribution of cancer deaths over time. We caution that HPVsim does not model cancer in detail (e.g. by using cancer stages). | logN(9,3) |
| **Immunity parameters** | | | |
| Initial B-cell-like immunity | imm_init | Initial B-cell-like immunity level assigned to women immediately following clearance. This is then used as a multiplier on the value of protective immunity they have to each genotype based on their history of infection/vaccination and the cross-immunity matrix. | Drawn from a beta distribution with mean of 0.35 and variance of 0.025, based on (7). |
| Immunity decay function | imm_decay | If provided, this determines the functional form of immunity decay over time | None (i.e. no decay) |
| Initial T-cell-like immunity | cell_imm_init | Initial T-cell immunity level assigned to women immediately following clearance. Used to derive protection against the probability of episomal infection transforming infected cells. | Drawn from a beta distribution with mean of 0.5 and variance of 0.025. No data were available to inform this, but we used a beta distribution to match how imm_init was modeled. |

| | | | |
|---|---|---|---|
| Immunity kinetics | imm_kin | If an immunity decay function is provided, the decay values over time are stored here | None by default; calculated automatically if a decay function is provided |
| Immune boost | imm_boost | If a person is re-exposed to a genotype with which they've already been infection, and subsequently clears that infection, this parameter determines the degree by which their pre-existing immunity levels will be boosted | 1 |
| Cross immunity matrix | immunity | Matrix containing entries that specify the degree of cross-immunity conferred by/to each genotype | See Table 1 |
| Medium cross immunity | cross_imm_med | Used to populate the cross-immunity matrix | See Table 1 |
| High cross immunity | cross_imm_high | Used to populate the cross-immunity matrix | See Table 1 |
| Probability of seroconversion | sero_prob | Probability of seroconversion | See Table S6 |
| **Genotype parameters** | | | |
| Genotypes | genotypes | List of genotypes to include in the simulation | HPV16, HPV18, and other high-risk (oncogenic) types, denoted as HRHPV |
| Genotype parameters | genotype_pars | All other genotype-specific parameters contained in Table S6 can be modified directly here. Default values are provided in Table S6, but alternatives can be specified here, e.g. `sim = hpv.Sim(genotype_pars = {16: {dur_episomal=4}})` | See Table S6 |
| **Other inputs** | | | |
| HIV parameters | hiv_pars | Parameters that determine the effect of HIV, if modeled | See Table S9 |
| Interventions | interventions | List of interventions to model; see Section 2.3 | |
| Analyzers | analyzers | List of analyzers to model | |
| Age bins | age_bins | List of age bins, used to determine by-age results | 0-5, 5-10, 10-15, 15-20, 20-25, 25-30, 30-35, 35-40, 40-45, 45-50, 50-55, 55-60, 60-65, 65-70, 70-75, 75-80, 80-85, 85+ |
| Standard population | standard_pop | Population distribution of the World Standard Population, used to calculate age-standardized rates (ASR) of incidence | SEER values (8) |

#### Table S3: HPVsim sexual network

| | |
|---|---|
| Relationship types | ‘default’ sexual network includes three relationship types: long-term, casual, and one-off; ‘random’ sexual network only includes a single relationship type capturing all relationships. |
| Relationship properties | Relationship duration; propensity for concurrency; coital frequency; condom usage; participation rates; and age mixing. |
| Network initialization | Relationship participation rates are used to determine the proportion of males and females in each age bin that are in relationships of each type. Each agent in the network is assigned a preferred number of simultaneous partners for each relationship type, governed by the concurrency propensity parameter for each partnership layer. Each female is assumed to begin with her preferred number of partners. Male partners are found using age mixing matrices, which specify the distributions of male partner ages for females of each specific age. Males are weighted higher for selection if they have fewer partners than their preferred number (e.g. 0 instead of 1), but can be selected even if they have a partner. Once both females and males have been selected, we assign each partnership a duration and a number of coital acts drawn from the partner type-specific distributions. Acts are scaled according to age and are assumed to increase between the age of sexual debut and the age of sexual peak, and then subsequently decline. |
| Network updating | At each time step (default = 0.25 years or 3 months), we dissolve relationships that have reached the end of their duration. For each relationship type and each age bucket, we calculate the difference between the proportion of females that are currently in relationships and the proportion assumed to participate in relationships of each type. We make up the difference by selecting females who are sexually active and weighting them higher for selection if they are under-partnered relative to their preferences. We then choose male partners by using the age mixing matrices, again weighting them higher for selection if they are under-partnered relative to their preferences. As with network initialization, relationships are then assigned a duration, condom usage, and an initial number of coital acts per year. |

### Figure S1: Sexual network for India

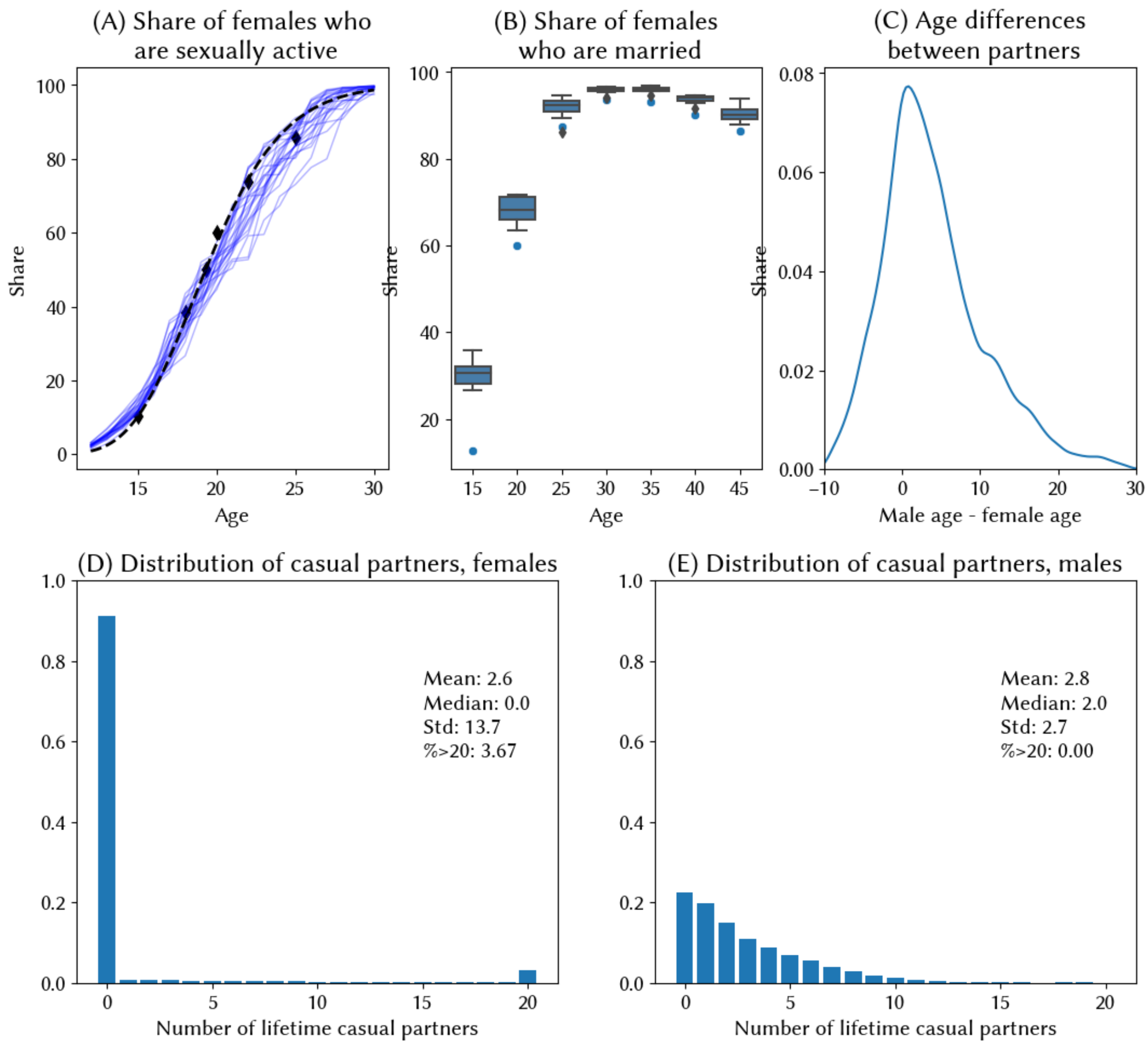


Figure S1: Metrics from HPVsim's sexual network model calibrated to data from India.

Panel A shows the share of females sexually active by age. Black diamonds represent the proportion of girls/women who report ever having personally experienced sexual intercourse by age 15, 18, 20, 22, and 25, and the median age at first sexual encounter. We treat these data points as quantiles of a lognormal distribution, and estimate the mean and standard deviation for each, which gives us an inferred distribution of the age of sexual debut (black dotted line). Finally, we input these inferred distributions into the model, and output the simulated proportion of females who ever had sex by age, following 20 cohorts of women over the course of their lives from age 0-30, with each cohort represented by a blue line.

Panel B shows the proportion of girls/women married by age 15-19, 20-24, 25-29, 30-34, 35-39, 40-44, and 45-49 (blue dots). We then find marriage incidence rates for each country to match these data points. The box plots summarize the simulated proportion of females in marriage-like relationships every 5 years measured from 1985-2020.

Panel C shows densities of the simulated differences between the ages of male and female sexual partners.

Panels D and E show the degree distributions for female and male casual partnerships, respectively.

### Table S4: Cross-immunity matrix following infection

| Protection[1] against ↓ after infection by → | HPV16 | HPV18 | hrHPV[2] |
|---|---|---|---|
| 16 | 1 | 0.5 | 0.3 |
| 18 | 0.5 | 1 | 0.3 |
| HR | 0.3 | 0.3 | 1 |

Table S4 notes:

1. An individual who clears infection and seroconverts is assigned a level of immunity to the genotype just cleared, $nab\_imm_{j,g}$, drawn from a beta distribution with mean of 0.35 and variance of 0.025, derived based upon a meta-analysis of natural acquired immunity (7). By default, this value stays constant over time, i.e. we do not model waning immunity, although simulations can be configured to include waning immunity with a specified decay function if desired (in which case $nab\_imm_{j,g}$ can be replaced by $nab\_imm_{j,g}(t)$). The individual *j*'s degree of protection against infection from any genotype $h$ is then calculated using the values in this cross-immunity matrix, as
$$inf\_imm_{j,h} = \sum_{i=1}^{G} \quad nab\_imm_{j,i} * ximm_{i,h}$$
where $ximm_{i,h}$ is the entry from the ith row and hth column of the cross-immunity matrix *ximm* in Table S4.
2. hrHPV: all other non-16/18 oncogenic HPV genotypes. Values are based on a compilation of evidence from cross-reactivity studies (9–11).

### Table S5: Prophylactic vaccine parameters

| | Bivalent[1] | Quadrivalent | nonavalent |
|---|---|---|---|
| 16 | 1 | 1 | 1 |
| 18 | 1 | 1 | 1 |
| HR | 0.5 | 0.5 | 1 |

Table S5 notes:

1. Cross-protection of the bivalent HPV vaccine against HPV31/33/45 and to a lesser extent 35 and 58 has been documented for up to 11 years as part of the Costa Rica HPV Vaccine Trial, with vaccine efficacy for 31/33/45 reported at 54.4% after one dose and 64.4% after 3 doses (12). Here, we assume 50% protection against these three genotypes (31/33/45) and lower levels (30%) against all other genotypes. We assume the same values for non-targeted genotypes for the quadrivalent and nonavalent vaccines.

#### Table S6: HPVsim genotype parameters and default values

| Type | Duration of infection prior to clearance or development of high-grade lesions (years)[1] | CIN function[2] | Duration of dysplasia prior to clearance or cancer (years) | Per-cell annual probability of transformation[2] | Relative transmissibility[3] | Probability of seroconversion after clearance of infection[4] |
|---|---|---|---|---|---|---|
| | dur_precin | cin_fn | dur_cin | transform_prob | rel_beta | sero_prob |
| 16 | logN(3,9) | Concave logistic function (see Eq1 in main paper) with k=0.3 | logN(5,20) | 0.002 | 1 | 0.75 |
| 18 | logN(2.5,9) | Concave logistic function (see Eq1 in main paper) with k=0.25 | logN(5,20) | 0.002 | 0.7 | 0.56 |
| HR | logN(2.5,9) | Concave logistic function (see Eq1 in main paper) with k=0.2 | logN(4.5,20) | 0.0015 | 0.6 | 0.60 |

Table S4 notes:

1. Chosen to fit to reported clearance/progression rates from Rodriguez [X]
2. Derived from calibration exercises.
3. Based upon a study by Brown et al (14) which reported the rate of seroconversion after HPV infection by type.

### Figure S2: Fitting the model to data from India

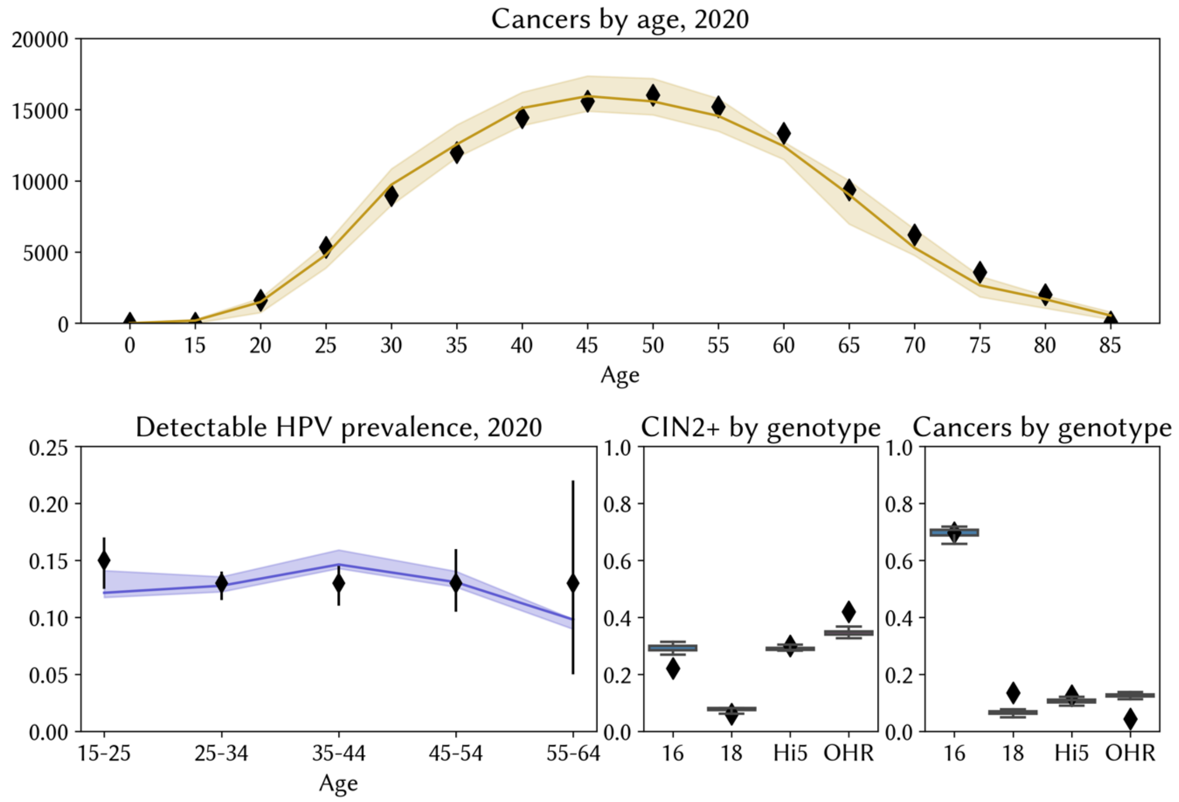


Figure S2 caption: HPVsim India model calibration. Data are shown as black diamonds and are sourced from Globocan 2020 estimates and HPV Information Centre compiled meta-analyses (15). We obtain a good fit to the distribution of cancers by age (top panel). In the bottom left panel, we show the fit to reported HPV prevalence estimates, although we note that this was not specifically included as a calibration target due to the significant variation in the quality of evidence and the difference in methodologies used across the studies from which these estimates were compiled. Our estimates of detectable HPV prevalence assume that among women with normal cervical cytology, 50% of infections acquired in a given year would be detected in that year. The model calibration could be notably improved with access to better HPV burden estimates. In the bottom right panels we show the modeled distribution of types found in women with high-grade CINs and cervical cancer (box plots) alongside estimates taken from Globocan/HPV Information Center.

### Figure S3: Estimates of cervical cancer burden in India

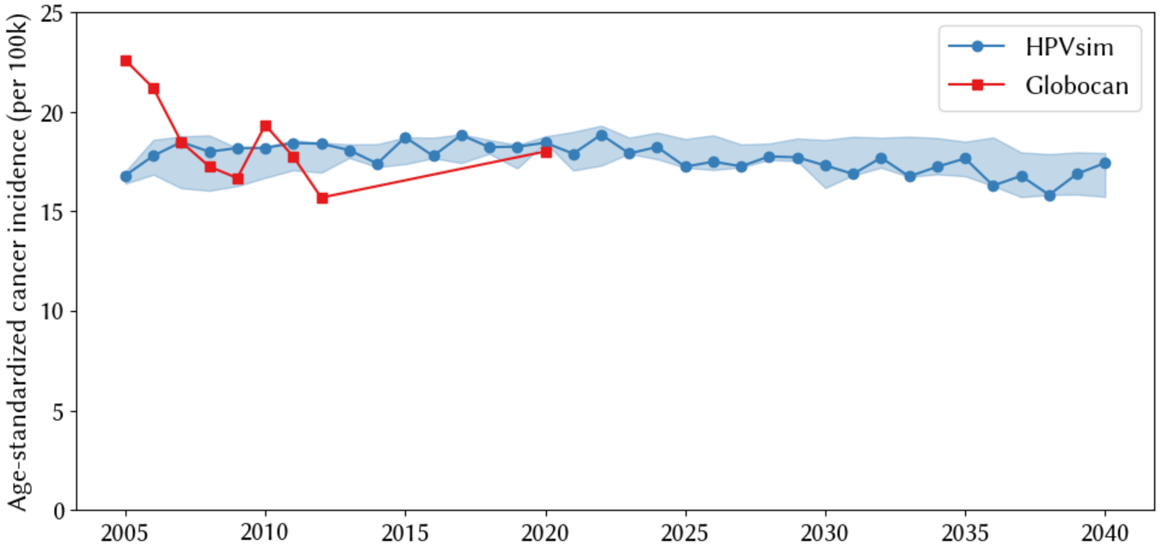


Figure S3 caption: Comparison of HPVsim and Globocan estimates of the age-standardized rate (ASR) of cervical cancer incidence in India.

### Table S7: Sensitivity/specificity of screening products

| | Visual inspection after acetic acid application (VIA)[1] | Liquid-based cytology (LBC)[2] | Conventional Pap test[3] | HPV[4] |
|---|---|---|---|---|
| Sensitivity for CINs[5] | 0.75 | 0.85 | 0.7 | 0.9 |
| Specificity[5] | 0.85 | 0.90 | 0.90 | 0.85 |

Table S7 notes:

1. We searched the literature for studies on VIA sensitivity/specificity. The most comprehensive estimates on sensitivity and specificity of all tests are from a 2016 meta-analysis by Mustafa et al. (16). Across studies that compared VIA to HPV DNA testing, the meta-analysis reports pooled estimates of VIA sensitivity and specificity of 69% (95% CI 54–81%) and 87% (79-92%). Across studies that compared VIA to cytology, VIA sensitivity and specificity are 77% (66%–85%) and 82% (67%–91%). Based on this, we use default assumptions of 75% sensitivity and 85% sensitivity, and assume that the test will identify 100% of carcinomas.
2. A Cochrane review of 40 studies and 140,000 women reported pooled sensitivity/specificity estimates for LBC of 72.9% and 90.3%, respectively (17). A study among >8000 women in Costa Rica found that LBC (using ThinPrep slides) detected 92.9% of cases with high grade squamous intraepithelial lesions (HSIL) or CIN2/3s and 100% of carcinoma cases (18). We assume that part of the reason for the different estimates of the sensitivity (72.9%(17) vs 92.9%(18)) is due to the fact that sensitivity differs depending on whether the true underlying state is CIN2 or CIN3. (The Cochrane review also notes that individual studies show large variation in reported sensitivity.) We therefore use default sensitivity values of 85% as the average of 80% for CIN2 and 90% for CIN3. For specificity, we use 90%, in line with the 90.3% reported in (17).

3. Mustafa et al report pooled estimates for conventional Pap smear sensitivity and specificity of 84% (76–90%) and 88% (79–93%) across studies comparing Pap smears to VIA, and 70% (57–80%) and 95% (92–97%) across studies comparing Pap smears to HPV testing. We also searched the literature to seek out estimates of sensitivity by disease stage (18–20). Based on this, we use default sensitivity values of 60% for CIN2 and 80% for CIN3 for the Pap smear. We use a default specificity of 90% (see note 5 on comparability to LBC).
4. Mustafa et al also report pooled estimates for HPV sensitivity and specificity as 94% (89–97%) and 88% (84–92%). We use default sensitivity of 90% (not varying by CIN grade) and default specificity of 85%. For HPV DNA test products, the sensitivity and specificity also vary depending on the HPV genotype. Our default assumptions assume that the HPV DNA test is targeted at genotypes 16, 18, 31, 33, 35, 45, 51, 52, 56, and 58. However, HPV assays can also return positive results when the sample contains a moderate to high viral load of HPV types not targeted by the probes in the assay (21). We therefore assume lower but non-zero sensitivity to all other genotypes.
5. Since our sensitivity and specificity values come from diverse sources across the literature, we need to ensure that the relative ranking of the tests is consistent with estimates from empirical studies comparing them. The Cochrane review described in note 3 found the relative sensitivity/specificity of HPV versus Pap for CIN 2+ to be 1.52/0.94, and versus LBC for CIN 2+ to be 1.18/0.96 (17). Our default assumptions therefore reflect the HPV test being more sensitive, but with lower sensitivity, than cytology (Pap or LBC). We also found two meta-analyses comparing conventional cytology to LBC, one which found no difference between the two (22), and one which found no difference in specificity but higher sensitivity of LBC compared to Pap (23). We assume slightly higher sensitivity for LBC but no difference in specificity. In line with the findings of Mustafa et al., our default assumptions imply that VIA is the least sensitive and least specific test.

### Table S8: Multi-scale modeling algorithm

1. Assume the "level 0" scale factor is 100 and the "level 1" scale factor is 1. In this case, one non-cancer agent corresponds to 100 people in the population and one cancer agent corresponds to one person in the population, but any numbers can be used: they do not have to be integers (although the ratio of these scale factors must be an integer; let's call it M). It's even mathematically valid for more than one cancer agent to correspond to a single person in the population (i.e., cancer agent scale factors of <1 are valid).
2. On a given timestep, assume N agents become infected with HPV. Without multiscale modeling, each of these N agents would be assigned a duration of infection and a disease outcome (such as clearance, dysplasia, or cancer). With multiscale modeling, a *matrix* of outcomes is generated instead from the same probability distribution, with N rows (corresponding to each level 0 agent) and M columns (corresponding to the additional cancer agents).
3. All agents (both level 0 and level 1) in the matrix who are scheduled to develop cancer are flagged.
4. Agents in the first column of the matrix who develop cancer will be kept as "level 0" agents for the purposes of their participation in disease transmission and demography, but are assigned the same scale factor (weight) as the cancer agents. For example, say that N=50 on this timestep, and 2 of these 50 agents are scheduled to develop cancer.
5. The remaining M-1 columns of the matrix correspond to potential new "cancer agents" who may be created. For example, say that in row 1 of the matrix, four of the M-1 = 99 columns

are flagged as cancer agents. Four new "level 1" (cancer) agents will be created. These new agents will inherit all the properties (e.g. age, sex) of the level 0 agent that represents this row, except with their individualized disease durations and outcomes calculated in step 2. This process is repeated for each row (i.e., each level 0 agent) in the matrix.

6. Once the new agents have been created, they are treated exactly the same as other agents in the model, with the exceptions that they do not participate in the sexual transmission network and are not counted in the calculation of crude birth rate.
7. When model results are calculated, outcomes are summed according to agent weight: in this example, a level 0 agent will be weighted 100 times higher in the prevalence calculation than a level 1 agent. However, since there are (on average) 100 times as many level 1 agents as there "should" be, the overall result is statistically correct.

### Table S9: HIV parameters

| **Parameter description** | **Default value** |
|---|---|
| Relative susceptibility to HPV acquisition (24) | 2.2 |
| Relative growth of HPV infection severity (24) | 0.5 |
| Relative risk of latent reactivation (24) | 3 |
